# Microbial and immune determinants of disease severity and death in pediatric pneumonia

**DOI:** 10.64898/2026.07.02.26356561

**Authors:** Karen L. Hanze Villavicencio, Ceylan Tanes, Clara Malekshahi, Daniel Cutillo, Maria Deloria Knoll, Christine Prosperi, Muge Kalaycioglu, Marlayna Harris, Paul J. Utz, Lisa M. Mattei, Daniel P. Beiting

**Affiliations:** Department of Pediatrics, University of Pennsylvania, Philadelphia, PA, USA; Division of Pediatric Infectious Diseases, Children’s Hospital of Philadelphia, Philadelphia, PA, USA; CHOP Microbiome Program, Division of Gastroenterology, Hepatology and Nutrition, Children’s Hospital of Philadelphia, Philadelphia, PA, USA; Department of International Health, Division of Global Disease Epidemiology and Control, John Hopkins University; Department of Medicine, Division of Immunology and Rheumatology, Stanford University School of Medicine, Stanford, CA, USA; Institute for Immunity, Transplantation and Infection, Stanford University School of Medicine, Stanford, CA, USA; Department of Pathobiology, School of Veterinary Medicine, University of Pennsylvania, Philadelphia, PA, USA

**Keywords:** Metagenome, Microbiome, Autoantibodies, Pneumonia, Pediatric, Upper Respiratory Tract, Sputum, Africa, Mali

## Abstract

Pneumonia is a leading cause of death globally and disproportionately affects children in lower- and middle-income countries. To explore microbial and immune correlates of disease and death, we performed metagenomic sequencing of upper respiratory tract (URT) microbiome in 309 children in Mali with pneumonia and 150 age- and season- and site-matched controls. We show that the URT microbiome matures throughout early life and is influenced by breastfeeding. URT microbiome maturation was disrupted during pneumonia resulting in loss of commensal species and expansion of pathobionts, which was linked to disease severity and death. Analysis of serum antibody levels revealed that low levels of passively acquired antibody from mothers, deficient antibody responses to RSV, and persistent autoantibody to cytokines were associated with pneumonia mortality in an age-dependent manner. These findings underscore the complex nature of pneumonia and identify microbial and immune factors for risk stratification and therapeutic interventions in pediatric pneumonia.

## Introduction

With an estimated global mortality rate of over 5 deaths per 1000 children, pneumonia ranks as the second leading cause of mortality among children under five years of age worldwide, responsible for 1 out of every 7 deaths^1^. Of the top 25 countries with the highest pediatric pneumonia mortality rates, 22 are in Africa^2^, underscoring the disproportionate impact of pneumonia on children living in low- and middle-income countries (LMICs)^3^. After the age of 5, pneumonia mortality rates plummet nearly 100-fold, demonstrating the extreme vulnerability of the youngest children to this disease^2,3^. The Pneumonia Etiology Research for Child Health (PERCH) study was a comprehensive multi-site case-control study conducted between 2011-2014 which aimed to estimate the causes of severe and very severe pneumonia among hospitalized children under 5 years of age from seven LMICs across Africa and Asia^4^ ^5^. Among HIV-uninfected cases, PERCH found that viruses accounted for the majority (61%) of severe pneumonia, with Respiratory Syncytial Virus (RSV) alone accounting for the largest etiological fraction of any pathogen (31%)^5^. Bacteria collectively accounted for 27% of cases, with *Mycobacterium tuberculosis, Streptococcus pneumoniae,* and the combined species of *Haemophilus influenzae* each accounting for approximately 6-7% of the etiological fraction^5^.

PERCH transformed our understanding of childhood pneumonia etiology in LMICs and shifted global health priorities toward prevention strategies aimed at reducing the burden of RSV-associated pneumonia and sustaining bacterial vaccination programs^6,7^. Indeed, despite having concluded over a decade ago, PERCH outcomes still guide World Health Organization (WHO) policies today, yet critical knowledge gaps remain. For example, PERCH used a combination of culture-dependent methods and multiplexed qPCR assay to detect pathogens previously linked to pneumonia – including 21 viruses, 1 fungus, and 11 bacteria (four at the genus level)^8–11^ – and did not employ untargeted sequencing to examine the broader species-level bacterial community present during pneumonia. As a result, there may be bacterial pathogens or virulence factors not targeted in the original study that are linked to disease. In addition, PERCH qPCR data for *Klebsiella pneumoniae* and *Moraxella catarrhalis* could not be used for etiology analysis due to poor assay specificity^5,8,12^. A recent postmortem study suggested that bacteria were responsible for more pediatric deaths from lower respiratory tract infections than viruses^13^, underscoring the need to capture a broader profile of bacteria associated with pneumonia in children. The microbiome includes many bacteria that are non-pathogenic, but which can dramatically influence pathogen virulence and the host inflammatory response. Notably, the microbiome is known to be highly dynamic in the first few years of life when it is strongly influenced by breast feeding^14,15^, raising the possibility that interpersonal differences in assembly of the microbiome in early life could influence pneumonia severity or outcomes. These limitations of PERCH prompted us to re-examine the PERCH specimens using metagenomic sequencing and immune profiling to capture a high-resolution view of the microbial and immune environment during pediatric pneumonia.

Although long considered a sterile environment, the lung is now recognized to be home to a low-biomass microbial community^16,17^ that develops dynamically in the first few months after birth^16,18^. Human studies using radio-labeled tracers administered to healthy volunteers^19^, together with microbial inoculation experiments in mice^20,21^, suggest that micro-aspirations occur commonly during sleep and result in an episodic flux of microbes moving from the high biomass environment of the upper respiratory tract (URT) to ‘seed’ the lung. Although rapidly cleared in a healthy lung, even a single aspiration event can impact lung immunity, for example decreasing susceptibility to pneumonia-causing pathogens, such as *S. pneumoniae*^21,22^. Conversely, multiple studies have linked decreases in URT microbiome diversity – resulting from viral infection^23^ or antibiotic exposure^24–26^ – to increased acquisition of lung pathobionts, including *Haemophilus influenzae*, *Moraxella catarrhalis*, and *Streptococcus pneumoniae*, and poor clinical outcomes ^22^.

Children under five years of age, particularly infants, are disproportionately susceptible to severe pneumonia owing to the immaturity of their immune system. Although conjugate vaccines represent one of the most impactful immunological interventions against childhood pneumonia, *Streptococcus pneumoniae* and *Haemophilus influenzae* continue to account for a substantial number of deaths annually among children under five^27,28^. In LMICs, this burden is driven not only by non-vaccine preventable serotypes and incomplete vaccine coverage but also by poorly characterized host immune factors that contribute to disease susceptibility and severity. The immunology of pediatric severe pneumonia encompasses multiple interrelated processes, including the developmental immaturity of innate and adaptive immunity^29^, prior pathogen-specific immune responses^30,31^, dysregulated inflammation that drives disease severity^32^, and host genetic and immune factors. While established risk factors such as malnutrition, HIV infection, and chronic comorbidities are well-recognized contributors to pneumonia mortality in LMICs^33^, the role of undiagnosed primary immunodeficiencies remains largely unquantified. Notably, between 1.3% and 10.5% of children with invasive pneumococcal disease – including those presenting with severe or recurrent pneumonia – are diagnosed with a primary immunodeficiency upon evaluation, most commonly involving antibody and complement deficiencies^34^. Taken together, these studies suggest that the URT microbiome and host immune responses may play an important role in shaping the outcome and severity of pneumonia but this needs to be explored in the most vulnerable patient population – young children living in LMICs.

## Results

### Age-dependent maturation of the URT microbiome during early life

We carried out detailed metagenomic and immune profiling of samples from 471 participants (321 cases with suspected severe or very severe pneumonia and 150 community controls without pneumonia) of the PERCH study, ranging in age from 1 to 59 months **(Supplemental Data 1)**. Our work focused specifically on Mali because this country experienced over 128 deaths per 1000 births in 2012 – the highest under-five mortality rate of all PERCH countries, and more than the next three top countries combined^2,5,35^. When the PERCH study began, Mali already had routine use of *Haemophilus influenzae* type b (HiB) vaccine and had just begun widespread use of 13-valent pneumococcal conjugate vaccine (PCV-13), yet still experienced the highest mortality rate within 30 days of hospital admission (12.9%)^5^, highlighting the importance of identifying additional immune or microbial factors that could be contributing to this devastating toll.

Nasopharyngeal/oropharyngeal (NP/OP) swabs and induced sputum are known to contain significant host contamination^36^ and relatively low microbial biomass, raising concerns that even deep metagenomic sequencing may yield limited insight into microbial community structure. To begin analysis of our metagenomic data, we filtered out human reads, removing a median of 77.9% and 83.7% of sequences from NP/OP and induced sputum, respectively. After host filtering, a median of 433,782 and 105,986 reads/sample were assigned to microbial taxa for NP/OP swabs and induced sputum, respectively. Microbiome profiling of low biomass samples is further complicated by contaminating microbial signal introduced from the environment and even molecular kit reagents^37–39^. We included extraction and library negative controls in our sequencing, which were comprised mainly of *Cutibacterium* and *Bradyrhizobium* genera **(Supplementary Figure 1A),** consistent with previous reports that these taxa are among the most common contaminants in microbiome studies^37–39^. The relative abundance of these two contaminants was two-orders of magnitude greater in our negative controls, compared to NP/OP swabs and induced sputum **(Supplementary Figure 1B)**. Setting a threshold of 2.7% combined relative abundance of the two contaminants and taxonomic assignment read count threshold of 10,000 resulted in the removal of 22/471 (4.7%) NP/OP and 46/265 (17.4%) induced sputum samples (**Supplementary Figure 1B and 1C),** resulting in 668 samples from 459 participants (309 cases and 150 controls) included in our analysis (**Supplemental Data 1)**.

**Figure 1:**
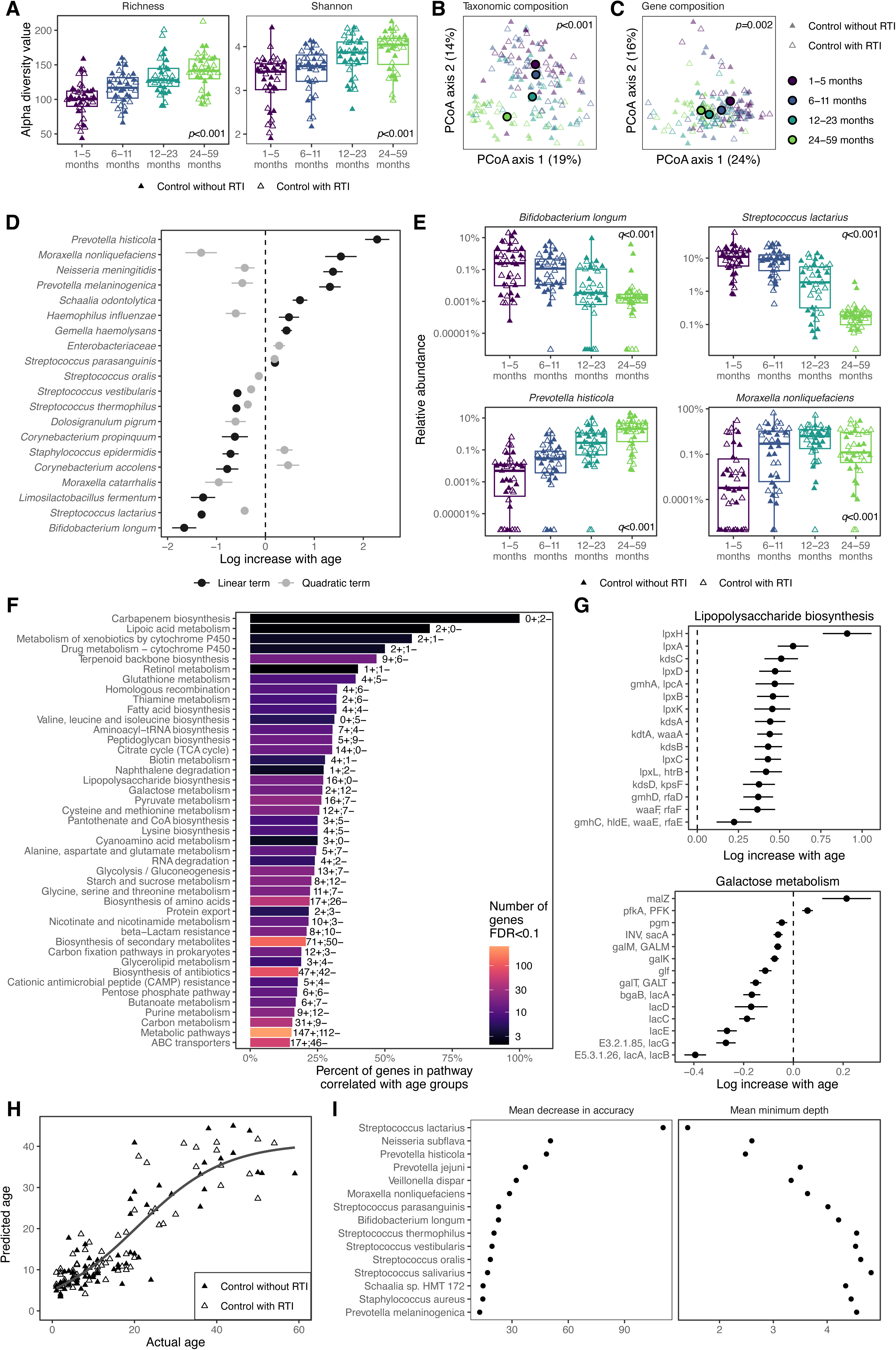
Age dependent maturation of the upper respiratory tract (URT) microbiome. **A)** Alpha diversity (number of unique species per 1000 reads and Shannon diversity) of samples over the four age groups. Linear models on age groups as ordered factors were used to test the changes in alpha diversity of the microbiome. Principal component analysis of Bray-Curtis distances of **B)** taxonomic composition and **C)** KEGG ortholog composition. The axis labels represent the percent variation captured by the axes. **D)** Species that are associated with age groups using linear models with the age group term modeled as an ordered factor. **E)** Relative abundance of two bacteria with the greatest positive and two with the greatest negative log change over age groups as seen in D. **F)** List of KEGG pathways that are enriched with orthologs that are associated with age groups using Fisher’s exact test. The x axis represents the percent number of genes in the pathway that are associated with age groups. Number of genes positively (+) and negatively (-) associated with age groups are shown next to each bar. **G)** Orthologs in the lipopolysaccharide biosynthesis and galactose metabolism that are associated with age groups. **H)** The actual age of the subjects versus the predicted age by using the maturity index. The maturity index was calculated using random forest. Line represents the self-starting logistic curve. **I)** The mean decrease in accuracy of the model when a feature’s values are randomly shuffled and the minimum depth of the feature averaged across all the trees built for the classification.

We first explored age-dependent changes in the URT microbiome. We stratified the controls into four age cohorts (1-5, 6-11, 12-23 and 24-59 months) and plotted the microbial diversity for NP/OP samples **(Figure 1)**. Both species richness and Shannon diversity showed marked increases with age **(Figure 1A),** indicating that the URT microbiome matures and diversifies throughout the first five years of life. Beta diversity analysis showed that both microbial community composition (**Figure 1B**) and gene content (**Figure 1C**) also differed significantly throughout early life, thus pointing to age-dependent maturation in the functional potential of the URT microbiome. PERCH included healthy community controls **(Figure 1, filled symbols)** as well as controls who had respiratory tract infection (RTI) but did not meet the criteria for pneumonia **(Figure 1, open symbols)**^40^. These latter controls allow us to distinguish between changes in the URT microbiome due to infection versus changes specifically linked to pneumonia. No significant differences in age-dependent URT microbiome maturation were observed between controls with and without RTI, indicating that URT maturation is a robust process.

Next, we sought to identify the microbial taxa and pathways driving this age-dependent maturation. Using linear models with the age groups as ordered factors, 20 bacterial taxa (19 identified at the species-level) showed either linear or non-linear (quadratic) association with age **(Figure 1D),** with *Prevotella histicola* and *Moraxella nonliquefaciens* showing the strongest positive association with age and *Bifidobacterium longum* and *Streptococcus lactarius* showing the strongest negative age association **(Figure 1D and 1E)**. Using Fisher’s exact test to find pathways enriched with genes associated with age, we identified 43 KEGG metabolic pathways **(Figure 1F)**, with all 16 genes involved in lipopolysaccharide (LPS) biosynthesis showing increased abundance with age **(Figure 1G, top).** In contrast, the abundance of genes involved in metabolism of lactose (lacA/B/C/D/E/G) and galactose (galM/K/T) decreased with age **(Figure 1G, bottom)**. Previous studies showed that stool microbiome could be used to predict chronological age^14^. Applying random forest regression to our taxon relative abundance data showed that, like the gut, the URT microbiome was a reliable predictor of actual age of a child **(Figure 1H)**; however, rather than many taxa contributing to the predictive strength^14^ we found that *S. lactarius* was an essential feature in the model, with a near complete loss of model accuracy if this single taxon was removed **(Figure 1I)**.

### Breast feeding shapes the URT microbiome

Diet has profound effects on the gut microbiome during health and disease, and breastfeeding in particular is linked to expansion of specific taxa that utilize human milk oligosaccharides as a substrate for growth^41^. The fact that 93.5% of our Mali cohort was breastfeeding during the first year of life **(Supplemental Data 1),** together with our observations that *B. longum* and *S. lactarius* – two taxa known to utilize breastmilk for energy – and metabolism of milk disaccharides (lactose and galactose) were strongly associated with age **(Figure 1D-G)**, prompted us to examine the role of breastfeeding in shaping the URT microbiome. Before 12 months of age, all but three participants were breastfed **(Figure 2A)**. Conversely, after 2 years of age, all but one participant in our cohort had transitioned off breast milk to solid food. However, 12 of the 36 participants in our 12–23-month group had been weaned while the others remained breast fed, and these two groups showed distinct microbial community structures **(Figure 2A)**. This provided a developmental window in which we could evaluate the impact of breast feeding on the URT microbiome. Linear models on log transformed bacterial abundances identified four age-associated taxa, including *S. lactarius* and three other *Streptococcus* species, that were significantly different between breastfed and non-breastfed infants at 12-23 months of age **(Figure 2B)**. These data show that age-associated maturation of the URT microbiome is partially influenced by breast feeding, and that these effects are most evident on *Streptococcus* species.

**Figure 2:**
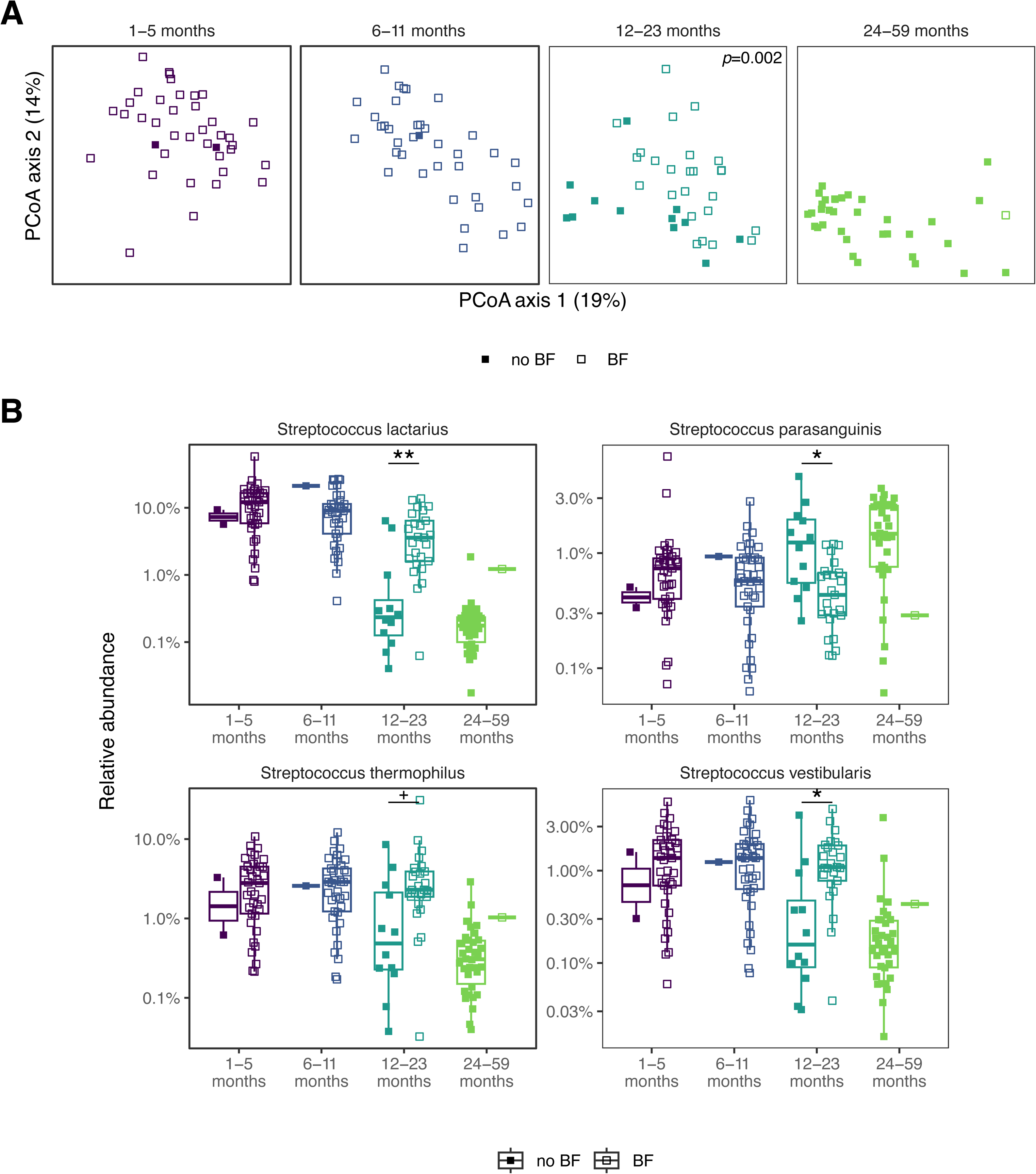
Effect of breastfeeding on the upper respiratory tract (URT) microbiome in PERCH controls. **A)** Principal component analysis of Bray-Curtis distances of taxonomic composition. The shapes represent of the subjects are breastfeeding at the time of sample collection. **B)** Relative abundance of bacteria that were associated with breastfeeding at the 12–23-month age group using linear models with the study group as a covariate and breastfeeding as the predictor. Asterisks represent FDR corrected p-values +<0.1, *<0.05, **<0.01, ***<0.001.

### Pneumonia disrupts the age-dependent maturation of the URT microbiome

We next explored how pneumonia impacts the maturation of the URT microbiome. We compared the NP/OP samples from the 150 controls (both with and without RTI) **(Figure 1)** to samples from 236 participants with suspected pneumonia who recovered from their illness, of which 150 were severe and 86 were very severe, as defined based on WHO classification at the time PERCH was conducted^40^. Compared to controls, children with pneumonia exhibited decreased URT microbiome diversity **(Figure 3A)**; increased dysbiosis index **(Figure 3B)**; and altered community structure **(Figure 3C)** and gene composition **(Figure 3D)** – all of which were most significant in very severe cases and children younger than 2 years. Moreover, this effect was not due to antibiotic use in the pneumonia cohort **(Figure 3A-D, point style)**. Several oropharyngeal commensals, including non-diphtheriae *Corynebacterium* species, *Veillonella atypica*, *Dolosigranulum pigrum*, and *Schaalia odontolytica* were significantly reduced in children with pneumonia, compared to healthy controls, particularly during early infancy (1-5 months)**(Figure 3E)**. Similarly, *S. lactarius* and other *Streptococcus* species – taxa that we showed were promoted by breast feeding **(Figure 2)** – were significantly reduced in children less than one year of age with pneumonia **(Figure 3F)**. The age-dependent relationship of *Moraxella nonliquefaciens* was also disrupted in children with pneumonia, most notably in children aged 6-11 and 12-23 months. When we examined the KEGG ortholog levels affected in pneumonia groups, we identified 16 genes altered in severe pneumonia compared to controls aged 1–5 months and 31 genes altered in children over 2 years. Similarly, we identified 321 genes altered only in very severe pneumonia compared to controls in the 6–11-month age group. Using Fisher exact test to identify pathways enriched with altered genes, we found several bacterial DNA/RNA repair and core metabolic pathways altered in very severe pneumonia compared to controls and 2−Oxocarboxylic acid metabolism and glycan degradation altered in severe pneumonia **(Figure 3G)**. The predominance of reduced gene abundance across multiple DNA repair and metabolic pathways suggests a loss of microbial functional diversity and depletion of metabolically versatile commensal taxa in children with very severe pneumonia. In addition to reduced commensals, we also detected increased *Enterobacteriaceae* in severe pneumonia cases aged 1-5 and 12-23 months, as well as in the very severe pneumonia cases over aged 2 years, with some cases having nearly 100% of the URT microbiome comprised of this family **(Figure 3F)**.

**Figure 3:**
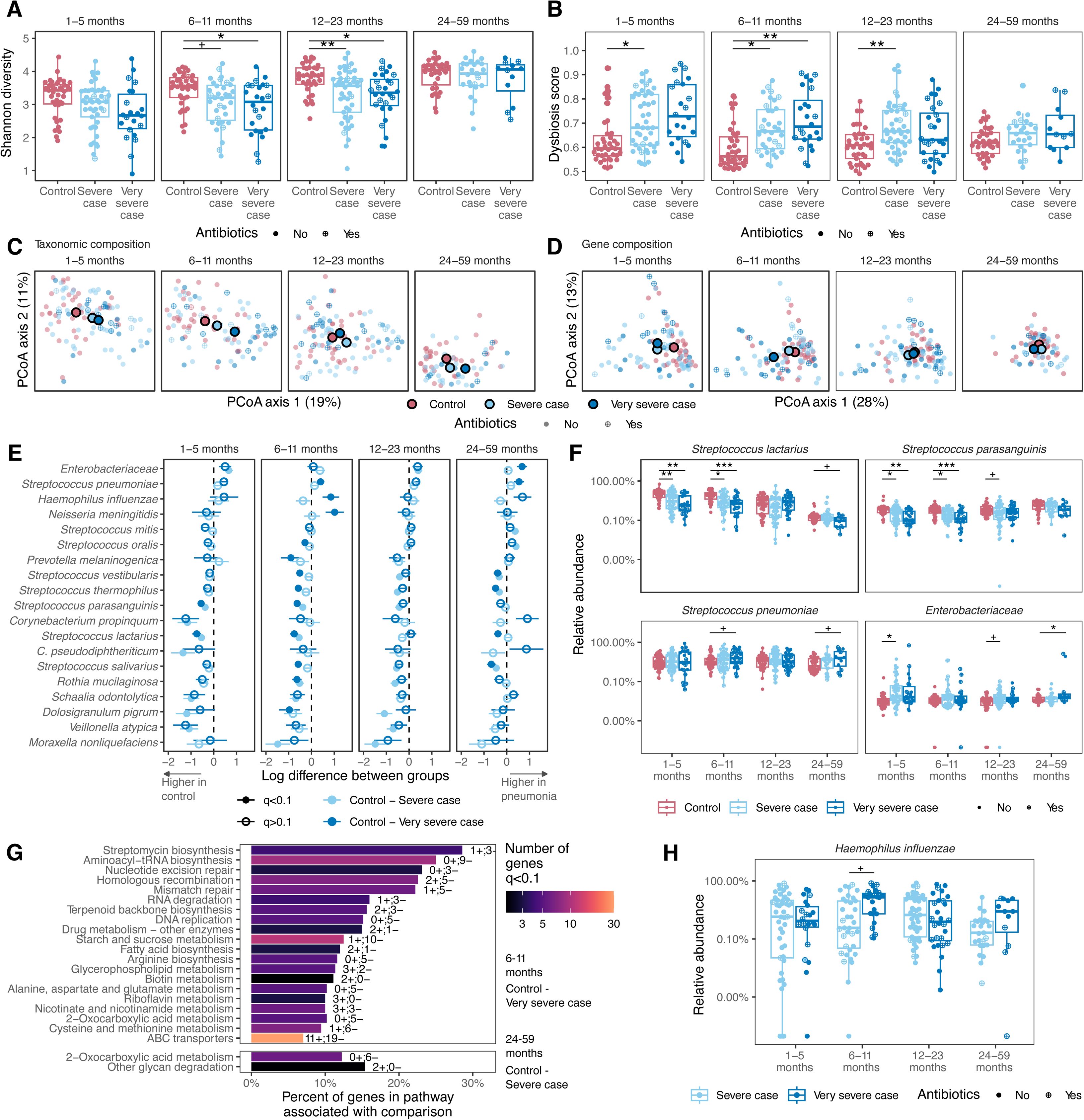
Changes in the upper respiratory tract microbiome (URT) during pneumonia. **A)** Shannon diversity of subjects across ge and study groups. **B)** Dysbiosis score of the samples calculated as the median Bray-Curtis distance from subjects in the control roup. Principal component analysis of **C)** taxonomic composition and **D)** KEGG ortholog composition. The axis labels represent the ercent variation captured by the axes. **E)** Log difference in relative abundance between pneumonia groups and the controls using inear models. Antibiotic use was used as a covariate. **F)** Relative abundance of two breastfeeding associated bacteria and two taxa that increase in subjects with pneumonia. **G)** List of KEGG pathways that are enriched with orthologs that are associated with pneumonia compared to controls. The numbers next to the bars represent the number of genes higher (+) and lower (-) in pneumonia roups. **H)** Relative abundance of Haemophilus influenzae in subject with severe and very severe pneumonia. Asterisks represent FDR corrected p-values +<0.1, *<0.05, **<0.01, ***<0.001.

Comparing severe and very severe cases identified limited but significant differences **(Figure 3H)**, with very severe cases having increased abundance of *Haemophilus influenzae* in children 6-11 months of age **(Figure 3G)**. To characterize differences between the URT and the lower respiratory tract microbiome, we then analyzed 219 induced sputum (IS) samples from severe and very severe pneumonia cases (IS was not collected from community controls). Even though we did not observe dramatic changes in the microbiome community in IS samples collected from severe and very severe subjects **(Supplemental Figure 2A)**, we identified lower levels of *Streptococcus* commensals and *Gemella haemolysans* during very severe pneumonia in the youngest age group independent of antibiotic exposure **(Supplemental Figure 2A and 2B).** Collectively, these data show that pneumonia is linked to the impaired development of the commensal community in the URT, which may create an opportunity for pathobionts to compete for resources. There were no microbial changes within the control group with and without RTI **(Figure 1)**, therefore the observed changes are specific to pneumonia and are not associated with respiratory tract infections more generally.

Several risk factors for pneumonia have been reported, including malnutrition, which was disproportionately observed in pneumonia cases, compared to controls, at all PERCH study sites^5^. Malnutrition has also been shown to delay the age-dependent maturation of the gut microbiome^14^. To test whether malnutrition might be an underlying factor driving the dysregulated URT microbiome in pneumonia, we used our random forest regression model derived from community controls **(Figure 1)** to assess the relative microbial maturation index (RMMI) in pneumonia cases **(Supplemental Figure 3)**. This data showed that, in contrast to what has been reported for the gut microbiome, the RMMI of the URT microbiome in malnourished controls is comparable to controls with normal nutritional status **(Supplemental Figure 3A and 3B)**. In children with pneumonia, the predicted microbiome age plateaued prematurely and diverged from chronological age, indicating a slightly higher maturation index **(Supplemental Figure 3A)**, but this was not correlated with malnutrition **(Supplemental Figure 3B).** Malnutrition had only a modest or no impact **(Supplemental Figure 3C)** on the four *Streptococcus* species that we identified as age-**(Figure 1)** and diet-dependent **(Figure 2)** URT commensals that are adversely impacted during pneumonia **(Figure 3).** Taken together with our results from Figure 2, these data implicate a dysregulated URT microbiome in both pneumonia severity and death, independent of malnourishment, antibiotics, and vaccination status **(Supplemental Figure 4)**.

### The URT Microbiome in Children Who Succumb to Pneumonia is Dysbiotic

Mali stands out amongst PERCH sites for high mortality rates^5,35^. To explore whether the dysregulated URT maturation we observed in pneumonia cases may be linked to death, we expanded our analysis to include an additional 63 participants with very severe pneumonia who died during hospital admission in PERCH **(Supplemental Data 1)**. NP/OP microbiomes from these cases revealed a dramatic reduction in microbial diversity **(Figure 4A)**; increased dysbiosis **(Figure 4B)**; and altered community structure **(Figure 4C)** and functional potential **(Figure 4D)**, compared to very severe participants who survived. These differences were statistically significant after 1 year of life and were marked by depletion of important commensals, including *Streptococcus* species, *Veillonella atypica*, *Schaalia odontolytica*, *Rothia mucilaginosa,* and *Gemella haemolysans, and Prevotella* species **(Figure 4E)**. Seven bacterial species increased in abundance in subjects who died in at least one age group **(Figure 4E, bold).** *Haemophilus influenzae* levels were consistently higher in children older than two who died **(Figure 4F)**. Given the prominent abundance of *H. influenzae* in fatal pneumonia cases **(Figure 4F)**, we used our metagenomic data to examine virulence factors by aligning the reads to the Virulence Factor Database. In children 24-59 months old, several virulence genes related to heme acquisition and utilization *- hemR, hxuA, hxuB and hxuC* – were associated with death from pneumonia with the greatest effect size **(Figure 4G and 4H)**. Together, these findings define a microbiome profile of fatal pneumonia in older children, characterized by loss of potentially protective commensals and an expansion of lung pathobionts with an enhanced array of virulence genes.

**Figure 4:**
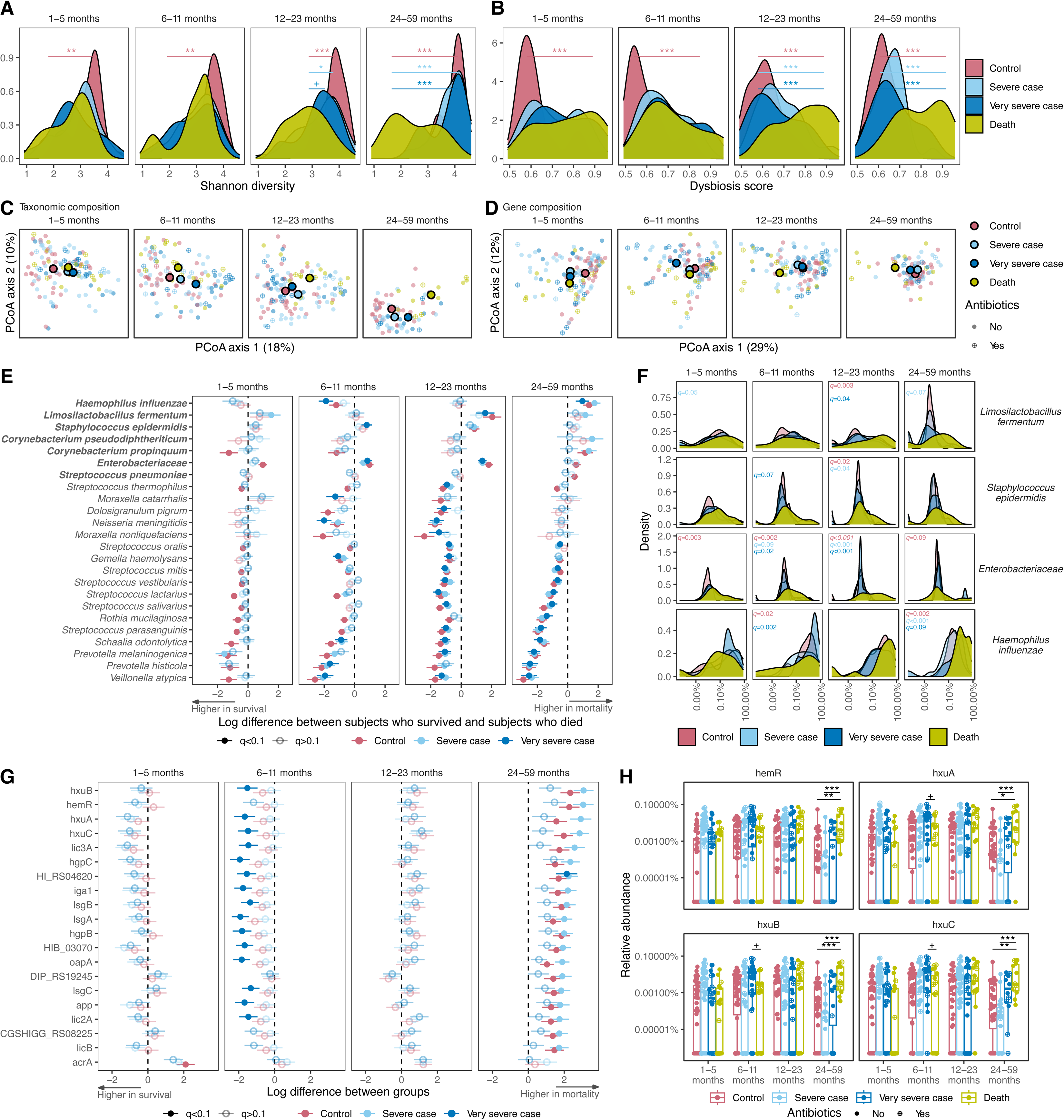
URT commensals are altered in pneumonia cases that die. Density plots of **A)** Shannon diversity and **B)** dysbiosis scores calculated as the median Bray-Curtis distance from subjects in the control group. The stars represent the results from linear model where we compare the groups where the subjects who survived to the subjects who died as a result of pneumonia. Principal component analysis of **C)** taxonomic composition and **D)** KEGG ortholog composition. The axis labels represent the percent variation captured by the axes. **E)** Log difference in relative abundance of taxa where subjects who died were compared to all the other groups using linear models. T he taxa that increased in abundance in at least one age group are bolded. **F)** Density plots for relative abundance of bacteria that are higher in subjects who died in at least one comparison based on results in E. **G)** Log difference in relative abundance of virulence factors where the subjects who died were compared to all the other groups using linear models. Only the top 20 virulence factors that are higher in subjects who died with the largest effect size are shown. **H)** Relative abundance of four virulence factors with the largest effect size as shown in G. Asterisks represent FDR corrected p-values +<0.1, *<0.05, **<0.01, ***<0.001.

### Pathogen exposure history is associated with pneumonia survival in children

One notable observation in the original PERCH study was that all pathogens detected in pneumonia cases were also found in controls, albeit usually to a lesser extent^5^, indicating that children are exposed to an array of viral and bacterial pathogens throughout early life. We reasoned that a child’s exposure history might influence the pneumonia outcomes. To explore this idea, we used a custom 73-plex autoantibody and 35-plex pathogen-antigen bead array to screen acute serum samples from our metagenomic cohort for both anti-cytokine autoantibodies and pathogen-specific antibodies.

We first examined the presence of pathogen-specific antibodies at the time of enrollment across all age groups. Except for SARS-CoV-2, which had not yet emerged as a pandemic virus at the time the PERCH study was conducted, all pathogen-specific antibodies on the array were detected above background in our cohort (see Methods) **(Figure 5A and Supplemental Data 2)**. Some antibodies tested showed relatively high average levels in the 1–5-month age group, a rapid decline by 6-11 months of age, and a rebound after 1 year of age, most notably those specific for Influenza and RSV antigens **(Figure 5A).** PERCH was conducted prior to the availability of RSV immunoprophylaxis and maternal vaccination, during a period when the highest burden of RSV disease occurred during the first year of life. These data therefore likely reflect a combination of early life exposure to pathogens, including RSV, and initial passive immunization of infants with maternal antibodies, which then wanes and is replaced by a *de novo* antibody response to pathogen exposure. Serum antibodies specific for Rhinovirus VP1 and RSV-F protein were the most abundant of all those tested and were already high at 6-11 months when other pathogen-specific antibodies were low, suggesting that exposure to these pathogens occurs very early in life **(Figure 5D)**.

**Figure 5:**
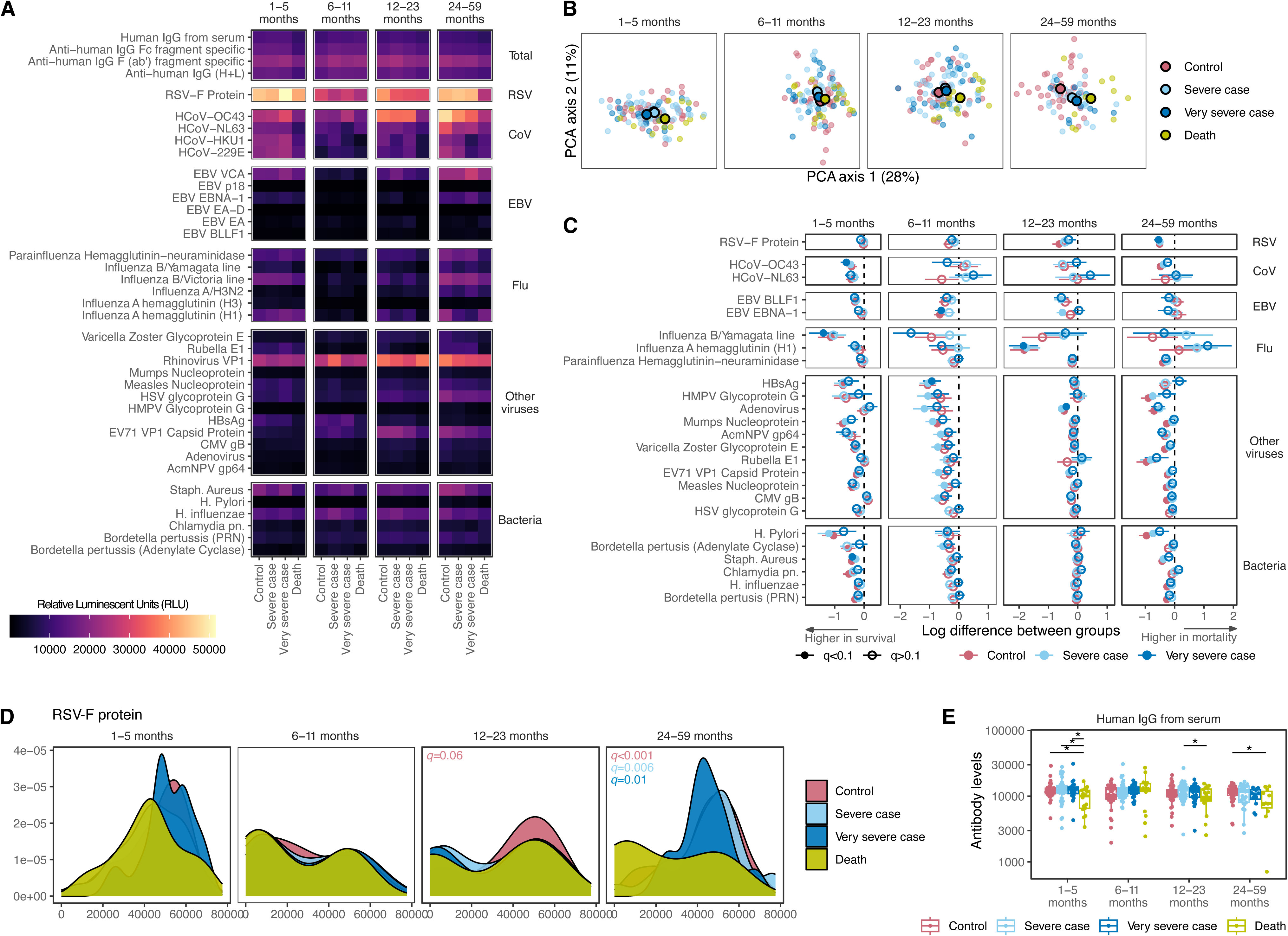
Pathogen-specific antibody profiles during pneumonia. A) Heatmap of average antibody levels against pathogens across age and study groups. B) Principal component plot of antibody levels against pathogens. The axes are labeled with the percent of variation captured by each axis. C) Log difference in levels of antibodies against pathogens where subjects who died were compared to all the other groups using linear models. D) Density plot of antibodies against RSV-F protein. E) Global IgG antibody levels. FDR corrected p-values are show. Asterisk indicates FDR < 0.05.

This age-dependent maturation of the antibody response from high maternal antibody in early infancy to high *de novo* pathogen-specific antibody later in childhood, was disrupted in children who died of pneumonia. This effect was most notable in the youngest and oldest age groups **(Figure 5B),** suggesting that both reduced passive immunization and reduced pathogen-specific antibody response are linked to pneumonia death. Of the 35 pathogen-specific antigens tested, 25 (71.4%) showed significant differences between children who died from pneumonia and any of the groups that survived (controls, severe, and very severe) **(Figure 5C)**. Stratifying these data by age group revealed specific differences in the immune status of PERCH participants who died from pneumonia. First, infants (1-5 months) who died showed significantly reduced antibody specific for influenza B, human coronavirus OC43, and *Staphylococcus aureus*, compared to very severe infants who survived **(Figure 5C, left, filled symbols)**. After 1 year of age, we still observed significantly reduced pathogen-specific antibody associated with mortality, but to different pathogens, including influenza A and adenovirus (12-23 months), and RSV (24-59 months) **(Figure 5C)**. Since 89% of the subjects were vaccinated against *H. influenzae* type b, the results are independent of vaccination status. The tendency for infants who died to have reduced pathogen-specific antibodies, compared to infants who survived pneumonia, prompted us to examine total serum IgG in our cohort. This analysis revealed significantly reduced total IgG in infants but not older children who succumbed to pneumonia **(Figure 5E)**, thus implicating ineffective passive immunization in infant pneumonia deaths.

These results prompted us to look more broadly at antibody specificity in our pneumonia cohort. Autoantibodies, especially anti-cytokine autoantibodies (ACAA), have emerged as a mechanism of disease pathogenesis that may predispose to infectious diseases^42^. Most recently, neutralizing autoantibodies to type-1 interferon have been associated with severe viral illnesses^43–45^. Therefore, we assessed the presence of 73 autoantibodies and ACAAs in our cohort using the same bead-based screening technology we used for anti-pathogen antibodies above **(Supplemental Data 2)**. Interestingly, autoantibodies in the control participants followed a trajectory during early life that was, broadly speaking, opposite from what we saw for pathogen-specific antibody, with autoantibodies declining with age in the control cohort **(Figure 6A).** Examining all 73 autoantibodies based on their abundance across the age cohorts identified 36 that differed significantly between controls and pneumonia cases, and which showed age-dependent autoantibody maturation **(Figure 6A)**. For example, at 6-11 months – an age when passive immunity is waning **(Figure 5A and 5D)** – nearly all controls had low but detectable ACAAs to IFN-alpha7, IFN-beta, and IFN-omega, and over half had autoantibodies to the IL1-family member, IL-33 **(Figure 6B, red bars).** In contrast, half or fewer of pneumonia cases who died had ACAA to these interferons, and only three had ACAA to IL-33 **(Figure 6B)**. This result suggests that low but detectable ACAA are part of the normal antibody repertoire and lower ACAA levels are associated with higher risk of death from pneumonia.

**Figure 6:**
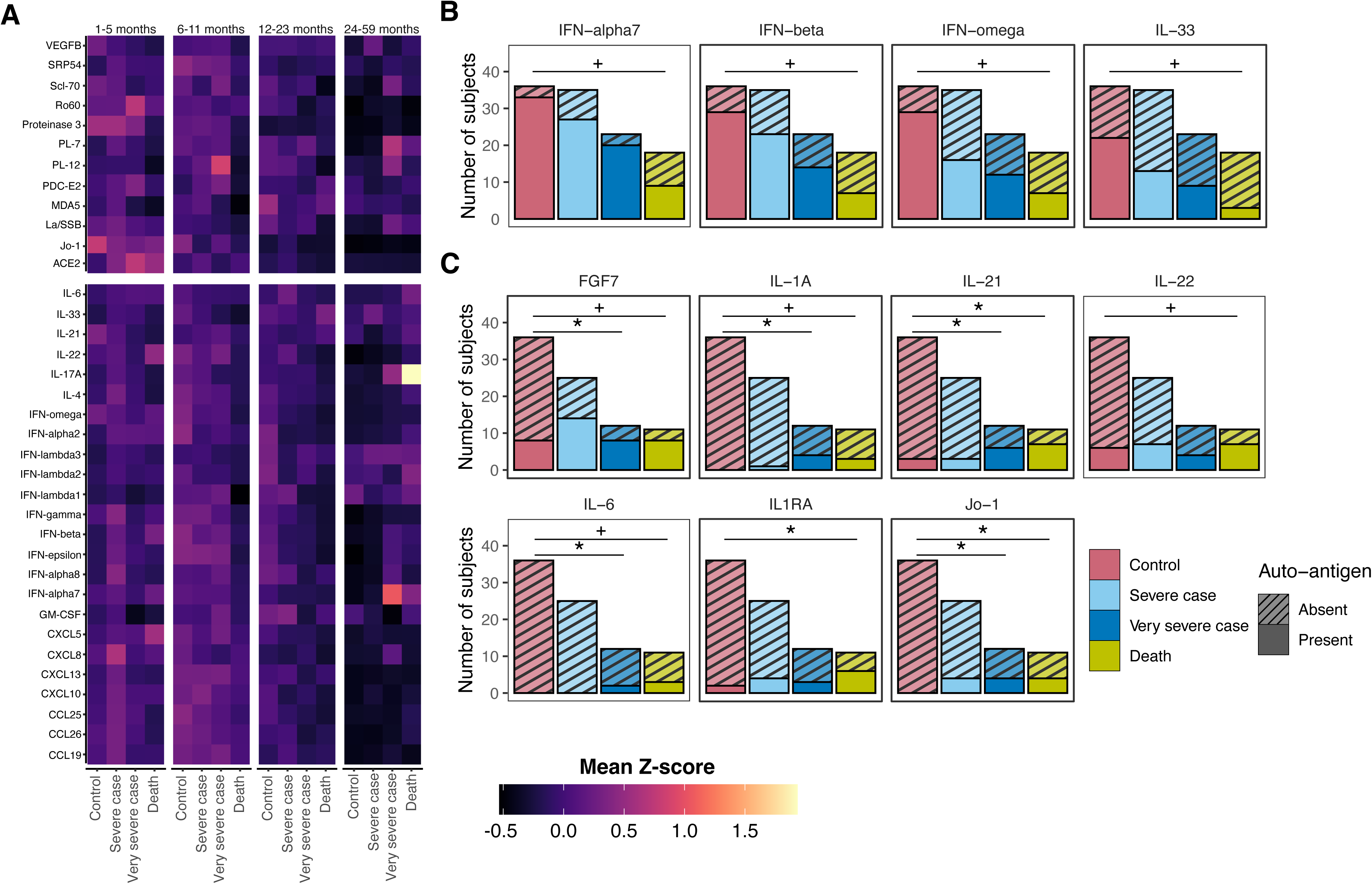
Serum autoantibodies during pneumonia. **A)** Heatmap of average autoantibody levels specific for 12 autoantigens (top) and 24 cytokines/chemokines (bottom) across age and study groups. Rows were clustered by Pearson correlation. **B-C)** Stacked bar plots showing number of participants positive (filled) and negative (striped) for autoantibodies that are statistically different by Fisher’s exact test comparing controls and death cohorts at **B)** 6-11 months and **C)** 24-59 months. Asterisks represent FDR corrected p-values +<0.1, *<0.05.

In contrast, by 24-59 months the opposite was true. As children aged and developed robust pathogen-specific antibody levels, autoantibodies declined **(Figure 6A)** and were either absent (IL-1A, IL-6, and Jo-1) or nearly absent (FGF-7, IL-21, IL-22, and IL-1RA) from control children **(Figure 6C)**. Strikingly, ACAA against IFN-alpha2 and IFN-alpha7 in children older than two years was associated with very severe pneumonia while autoantibodies to IL-17A were prevalent in children who died from pneumonia, compared with controls **(Figure 6A, far right column).** Although insufficient serum volumes were available for receptor blocking studies, inhibitory ACAA against these soluble factors have been shown to be pathogenic in COVID-19 and other respiratory infections^46–49^. Taken together with our anti-pathogen antibody data, these results point to a dynamic immunological process that unfolds in early life in which both passive immunity and autoantibodies decline from birth to age five, while anti-pathogen titers rise with exposure to viral and bacterial microbes. Although it is unclear how, this maturation is disrupted in some children leading to impaired pathogen-specific antibody responses and elevated autoantibodies, which are risk factors for severe pneumonia and death.

## Discussion

Pneumonia is a complex and multi-factorial disease with co-infections and environmental factors known to be risk factors for severe disease. Moreover, URT commensal microbes episodically transit to the lung^21^, underscoring the importance of examining this complex microbial community during pneumonia, particularly in children from Africa, where early life pneumonia mortality rates are some of the highest in the world. Our metagenomic analysis of the URT microbiome and induced sputum from 459 children under the age of 5 living in Mali – including 150 controls and 309 pneumonia cases – provides a detailed view of respiratory tract microbiome development in early life and its potential role in pneumonia severity and mortality. We found that the URT microbiome matures throughout early life **(Figure 1)** and is heavily influenced by breastfeeding **(Figure 2)**. Using shotgun metagenomics, we identified *Streptococcus lactarius* as the dominant taxa linked to breastfeeding and driving age-associated microbiome maturation in our cohort. Originally identified in human breast milk, *S. lactarius* displays unique metabolic capabilities, including fermentation of lactose and other related carbohydrates supporting its adaptation to the breast milk environment^50^. Notably, its abundance in breast milk peaks during the first six months of breastfeeding and declines with the introduction of solids, mirroring the pattern we observed in the URT^41^. Seminal studies examining the gut microbiome in children in Bangladesh showed a very similar age-dependent maturation and revealed that dietary or microbial therapeutics were able to modulate maturation to ameliorate disease^14,51,52^. Thus, our data raise the possibility that the URT microbiome is similarly involved in respiratory disease and that the microbial environment and immune state of the lung might be amenable to manipulation by targeting the URT microbiome, for example with oral prebiotics or probiotics.

We observed reduced URT microbiome diversity during pneumonia, marked by a loss of commensal taxa, most notably *Prevotella melaninogenica*, *Corynebacterium propinquum* and *pseudodiphtheriticum*, *Schaalia odontolytica*, *Moraxella nonliquefaciens*, *Veillonella atypica*, and Dolosigranulum pigrum **(Figure 3)**. Several of these species have been previously shown to inhibit airway pathobionts. For example, *P. melaninogenica* has been associated with rapid clearance and protection against *S. pneumoniae* infection in murine models by inducing TLR2-dependent neutrophil production of TNF-α^22^; *Corynebacterium species* can inhibit the growth of *S. pneumoniae* in the nasopharynx of children by releasing free-fatty acids^53^; and *D. pigrum* is consistently associated with reduced *Staphylococcus aureus* and *S. pneumoniae* nasopharynx colonization and lower respiratory tract infection in early infancy^54^. Given these findings, we hypothesize that reduced abundance of multiple protective URT commensal species during pneumonia leads to a loss of colonization resistance against pathobionts, a phenomenon that has also been reported in the gut with Clostridia^55,56^ and that is attributed to metabolic competition between closely related species^57^. Our finding establish only associations, and additional mechanistic animal studies employing adoptive transfer of microbial communities and defined consortia are needed to formally establish causality between URM microbes and protection from pneumonia.

The PERCH study was critical in defining the viral and bacterial pathogens driving pneumonia in LMICs, yet many questions remained after the conclusion of the study in 2014. Despite the extensive molecular diagnostic work in PERCH^8^, no differences in the number or type of pathogens were detected in children based on disease severity or mortality^5^, highlighting the need to more broadly profile the microbial communities present during pediatric pneumonia, and to do so at species-level resolution^22^. We identified *H. influenzae* and *Enterobacteriaceae* as pathobionts associated with very severe pneumonia and death. *H. influenzae* commonly colonizes the URT asymptomatically but can transition to invasive disease when the surrounding microbial and host environment is perturbed^23^. Consistent with the breakdown of colonization resistance described above, carriage of the commensal bacterium *Haemophilus haemolyticus* has been associated with decreased non-typeable *H. influenzae* colonization, suggesting that closely related commensals may constrain this known pneumonia pathogen^58^. Notably, we observed the highest levels of *H. influenzae* in patients with the worst outcomes (very severe disease and death), which was associated with increased levels of virulence factors responsible for heme acquisition and utilization (*hemR, hxuA, hxuB and hxuC*). Together, these data support a model in which URT dysbiosis and virulence gene enrichment act synergistically, facilitating expansion of invasive *H. influenzae* strains, and contributing to poor clinical outcomes.

The relationship between the ‘immunological history’ of early life pathogen exposure and resistance to subsequent infections is complex. Not surprisingly, studies of RSV in birth cohorts have shown that prior RSV infection and high pre-existing RSV antibodies have been associated with lower risk of RSV illness in children^59^, however this protection may be transient and require repeated exposures to mount robust and durable protection^60^. In contrast, cross-pathogen protection may induce a broader non-specific innate immune state leading to enhanced proinflammatory responses upon subsequent re-exposures^61,62^. In our cohort we found that infants who had reduced pathogen-specific antibodies had poorer outcomes suggesting suboptimal immune maturation in early life or inadequate passive immunity. Passive immunity through transplacental transfer of IgG during pregnancy and IgA in breast milk can be influenced by maternal nutritional and immune status. During the study period in Mali, high rates of malnutrition were observed in children, which may also reflect maternal nutritional deficiencies.

In addition to microbial and host immune factors, underlying immunologic abnormalities may increase susceptibility to severe infections. Anti-cytokine autoantibodies (ACAA) have emerged as one potential cause of acquired immunodeficiency, as patients with ACAA can exhibit clinical phenotypes resembling individuals with pathogenic mutations predisposing to infections^63^. Importantly, ACAA are not always associated with disease and have been detected at low levels in healthy individuals^42^. Seroprevalence studies further indicate that ACAA are frequently observed following acute illness, such as in acute respiratory distress syndrome, severe COVID-19 and sepsis^44,45,64^, and decline in convalescence following the initial infection^65^. This may potentially reflect a shift from a proinflammatory to an anti-inflammatory state aimed at limiting tissue damage and preventing excessive immune activation following the initial infection^65^. In our cohort, we identified a diverse array of ACAA targeting pathways involved in inflammation and tissue repair both in healthy controls and pneumonia groups. Notably, persistent ACAA were associated with pneumonia mortality in an age-dependent manner with a higher prevalence of ACAA in children older than two years who had very severe pneumonia (ACAA directed against Type I IFNs, IFN-alpha2 and IFN-alpha7) or who died from pneumonia (ACAA recognizing IL-17A) compared with controls. This suggests that specific ACAA may play a role in infections beyond SARS-CoV2 and influenza^46–49^. The precise role of ACAA in the pathogenesis of pediatric pneumonia, as well as their clinical implications, warrant further investigation.

## Materials and Methods

### PERCH cohort and pneumonia definition

All samples were collected as part of the original PERCH study conducted between January 2012 to January 2014 at l’Hôpital Gabriel Touré in Bamako, Mali, which was approved by the Institutional Review Boards (IRB) or Ethical Review Committees for each participating institution and at The Johns Hopkins Bloomberg School of Public Health. Use of de-identified demographic and clinical data study was approved by the IRB of the University of Pennsylvania (IRB # 854435). Samples were obtained through close coordination with the PERCH Core Team at Johns Hopkins University. Ethical approval for this study was obtained from l’Hôpital Gabriel Touré in Bamako, Mali (reference # 00000138).

Cases and controls were defined as described previously in the PERCH study^5,40^. Briefly, eligible cases were those admitted to hospital meeting the 2005 WHO definition of severe or very severe pneumonia. “Severe pneumonia” was defined as cough or difficulty breathing with lower chest wall indrawing. “Very severe pneumonia” was defined as cough or difficulty breathing and at least one of the following signs: central cyanosis, difficulty breastfeeding or drinking, vomiting, convulsions, lethargy, unconsciousness, or head nodding. Community controls were age- and season-matched to cases and randomly selected from the same catchment area as cases regardless of respiratory symptoms, excluding those who met the criteria for PERCH case enrollment. Detailed demographic, environmental, and clinical characteristics were collected from cases and controls, including vaccination status, breastfeeding practices, nutritional status, antibiotic exposure, respiratory support and outcome. All PERCH clinical and epidemiological data are available on https://ClinEpiDB.org^66^.

Nasopharyngeal (NP) and oropharyngeal (OP) swabs were collected in all cases and controls at the time of enrollment, ideally before starting antibiotics, as previously described^8,67^. The NP swab was collected first, followed immediately by the OP swab. Both swabs were placed into the same vial containing 3 ml of universal transport medium (UTM) (Copan), vortexed for 20-30 seconds, and then discarded. Induced sputum (IS) was only collected from cases, not controls, and was obtained within 48 hours of hospital admission, by trained medical staff through the induction of deep coughing by inhalation of hypertonic saline. Blood for acute serum from both cases and controls was also obtained at recruitment. Antibiotic administration and activity levels in serum were measured upon enrollment in all participants. Aliquots of NP/OP UTM, induced sputum, and acute serum were stored at -70°C at the study specimen biorepository (Canterbury Health Laboratories, Christchurch, New Zealand)^9^. All samples were shipped on dry ice to the University of Pennsylvania School of Veterinary Medicine in Philadelphia, where they were stored at -80°C until processing.

Of the original 674 cases and 725 controls enrolled at the Mali site during the original PERCH study, 471 were selected for this study, to obtain a 2:1 ratio of cases to controls and a balanced representation of four age groups: 1-5, 6-11, 12-23 and 24-59 months **(Supplemental Data 1)**. Cases were selected only if they had paired upper (i.e., NP/OP) and lower (i.e., IS) respiratory tract specimens and complete clinical data.

### Metagenomic sequencing

DNA was extracted from IS and NP/OP swab samples using the DNeasy PowerSoil Pro kit (QIAGEN) on QIAcube robots. Libraries were prepared from up to 10 ng of extracted DNA using the Illumina DNA Prep kit and IDT for Illumina unique dual indexes. Negative extraction controls (reagents only) and negative library controls (microbial DNA-free water) were processed alongside the NP/OP and IS samples to assess background contamination from reagents and the environment. Sequencing libraries were combined into two separate pools, which were sequenced on the Illumina MiniSeq to evaluate quality and barcode distribution. Each pool was then sequenced on two lanes of an S4 300-cycle kit using an Illumina NovaSeq 6000 instrument (Center for Applied Genomics, Children’s Hospital of Philadelphia).

Shotgun metagenomic data were analyzed using Sunbeam v.4.6.0^68^. Adapter and low-quality sequences were removed, and human-derived reads were discarded as part of quality control steps using default parameters of the pipeline. The abundance of bacteria was estimated using Kraken^69^ with a database of genomes downloaded June 2024. Reads were mapped to Kyoto Encyclopedia of Genes and Genomes (KEGG) protein database^70^ and Virulence Factor Database (VFDB)^71^ using Diamond search^72^ to estimate the abundance of bacterial gene orthologs and bacterial virulence factors, respectively.

### Serum antibody profiling

Acute sera stored at -80°C were thawed and analyzed using a previously described multiplexed bead array^73^. In brief, the array was constructed by coupling antigens to carboxylated magnetic beads with unique barcodes, which were then distributed to 96-well plates and mixed with a 1:100 dilution of heat-inactivated patient serum in duplicate. After incubation and washing, beads were labeled with a fluorescently conjugated secondary antibody and then analyzed on a Luminex FlexMap3D^TM^ instrument with Luminex xPONENT^®^ version 4.2 software. A minimum of 100 events per bead ID were counted, and binding events were displayed as Mean Fluorescence Intensity (MFI). All data analysis and statistics were performed using R/Bioconductor. For normalization, average MFI values for “bare bead” IDs were subtracted from average MFI values for antigen conjugated bead IDs. The average MFI for each antigen was calculated using samples from the healthy cohort. Antibodies were considered positive if MFI was ≥ 3000 units and ≥ 5 SD above the average MFI for the healthy controls for that antigen.

### Statistical analysis

*Cutibacterium* and *Bradyrhizobium* genera were defined as contaminants based on their high prevalence in our negative control samples in addition to being well-documented as two of the most common contaminants in low-biomass microbiome studies^37–39^. We excluded from the analysis 68 samples that had a total relative abundance of these genera that was ≥ 2.7% (two standard deviations lower than the mean abundance) or less than 10000 reads assigned using Kraken. R software version 4.4.0 and RStudio software version 2024.04.0+735 were used for data analysis and visualization. Microbial community-level differences between groups were assessed with Permutation Multivariate Analysis of Variance (PERMANOVA) on Bray-Curtis distances. Linear models were used to identify associations with either bacterial abundance, gene abundance, alpha diversity, dysbiosis score, or antibody levels as outcome and study groups, age groups or breastfeeding status as predictors. Antibiotic use was added as a covariate. Only the taxa with more than 1% mean relative abundance in at least one study group were tested. One-tenth of the smallest non-zero relative abundance was added to the values and log10 transformed before testing. We grouped the KEGG orthologs that were statistically significant in a comparison into pathways and identified the pathways that are enriched using Fisher’s exact test. We considered *p*-value ≤ 0.05 to be statistically significant. When multiple comparisons were made, *p*-values were corrected using Benjamini-Hochberg to generate false discovery rates (FDR) and only features with FDR ≤ 0.1 were considered. To calculate the maturity index, a random forest model was built with the ages as outcome and the relative abundance of taxa with at least 0.5% mean across the NP/OP samples of healthy, non-malnourished subjects as predictors. 10,000 trees were built using a third of the features selected randomly for each tree. Mean minimum depth of the features was calculated using randomForestExplainer package^74^.

## Data availability

Raw Illumina sequencing reads for PERCH microbiome data are available on the Sequence Read Archive (SRA) accession, PRJNA1474087). Processed microbiome data are publicly available for data mining, analysis, and visualization on https://MicrobiomeDB.org^75^. Clinical and epidemiologic data on PERCH participants are available in https://ClinEpiDB.org^66^. Luminex antibody panel data are available in **Supplemental Data 2.**

## Code availability

The code used to generate all figures and analyses can be found on GitHub at https://github.com/ctanes/perch_metagenome.git.

## Contributions

Planning and project management: K.L.H.V., C.T., L.M.M., D.P.B. Sample organization and management: C.M., D.C., K.L.H.V., L.M.M. Serum antibody assays: M.H., M.K., and P.J.U. Planning and advising on analysis and interpretation of clinical and epidemiological data from PERCH: M.D.K. and C.P. Data analysis: C.T., K.L.H.V., L.M.M., D.P.B. Computational resource management: C.T. Writing of the manuscript: C.T., K.L.H.V., L.M.M., D.P.B.

## Funding

This research was supported by pilot grants from the PennCHOP Microbiome Center and the University Research Foundation fund (URF) at the University of Pennsylvania. K.L.H.V. was supported by the St. Jude-PIDS Fellowship Award in Basic and Translational Science and the Research in Academic Pediatrics Initiative on Development (RAPID) Scholars Award. PJU was also supported by the Henry Gustav Floren Trust; Stanford Department of Medicine Team Science Program; Stanford Medicine Office of the Dean; NIAID R01AI175771 and R01AI182319-02. The PERCH study was supported by grant 48968 from the Bill & Melinda Gates Foundation to the International Vaccine Access Center, Department of International Health, Johns Hopkins Bloomberg School of Public Health (Baltimore, MD, USA). Sample shipping from the PERCH biorepository in New Zealand was supported by the Bill and Melinda Gates Foundation (Investment ID INV-052361 and INV-067813). M.D.K. and C.P. received no additional support for this analysis.

## Acknowledgements

We thank the ClinEpiDB team (Drs. Danica Helb, Sheena Tomko, and Nupur Kittur) for input on study design and accessing clinical and epidemiological data on https://ClinEpiDB.org. We thank Sierra Brodhage and Maya Mathur for assistance with scanning and organizing PERCH samples and inventory. We thank Drs. Rui Xiao and Andrew Hart for assistance with figure design.

## Notes

### Competing Interest Statement

The authors have declared no competing interest.

### Author Declarations

IRB of the University of Pennsylvania gave eithical approval for this work (IRB # 854435). Ethics committee of Gabriel Toure Hospital in Bamako, Mali gave ethical approval for this work (reference # 00000138).

## References

1. Perin, J. et al. Global, regional, and national causes of under-5 mortality in 2000–19: an updated systematic analysis with implications for the Sustainable Development Goals. Lancet Child Adolesc. Health 6, 106–115 (2022).

2. Villavicencio, F. et al. Global, regional, and national causes of death in children and adolescents younger than 20 years: an open data portal with estimates for 2000-21. Lancet Glob. Health 12, e16–e17 (2024).

3. Bender, R. G. et al. Global, regional, and national incidence and mortality burden of non-COVID-19 lower respiratory infections and aetiologies, 1990–2021: a systematic analysis from the Global Burden of Disease Study 2021. Lancet Infect. Dis. 24, 974–1002 (2024).

4. Levine, O. S. et al. The Pneumonia Etiology Research for Child Health Project: a 21st century childhood pneumonia etiology study. Clinical Infectious Diseases 54 **Suppl 2**, S93–101 (2012).

5. O’Brien, K. L. et al. Causes of severe pneumonia requiring hospital admission in children without HIV infection from Africa and Asia: the PERCH multi-country case-control study. The Lancet 394, 757–779 (2019).

6. Duke, T. What the PERCH study means for future pneumonia strategies. The Lancet 394, 714–716 (2019).

7. Klugman, K. P. & Rodgers, G. L. PERCH in Perspective: What Can It Teach Us About Pneumonia Etiology in Children? Clin. Infect. Dis. 64, S185 (2017).

8. Driscoll, A. J. et al. Standardization of Laboratory Methods for the PERCH Study. Clin. Infect. Dis. 64, S245 (2017).

9. Murdoch, D. R. et al. Laboratory methods for determining pneumonia etiology in children. Clinical Infectious Diseases 54 **Suppl 2**, S146–52 (2012).

10. Driscoll, A. J. et al. Evaluation of fast-track diagnostics and TaqMan array card real-time PCR assays for the detection of respiratory pathogens. J. Microbiol. Methods 107, 222–226 (2014).

11. Cissé, A. et al. Comparison of performance between Fast Track Diagnostics Respiratory Kit and the CDC global reference laboratory for influenza rRT-PCR panel for detection of influenza A and influenza B. Influenza Other Respir. Viruses 15, 381–388 (2021).

12. Zar, H. J. et al. Aetiology of childhood pneumonia in a well vaccinated South African birth cohort: A nested case-control study of the Drakenstein Child Health Study. Lancet Respir. Med. 4, 463–472 (2016).

13. Bassat, Q. et al. Causes of Death Among Infants and Children in the Child Health and Mortality Prevention Surveillance (CHAMPS) Network. JAMA Netw. Open 6, E2322494 (2023).

14. Subramanian, S. et al. Persistent gut microbiota immaturity in malnourished Bangladeshi children. Nature 2014 510:7505 510, 417–421 (2014).

15. Shenhav, L. et al. Microbial colonization programs are structured by breastfeeding and guide healthy respiratory development. Cell 187, 5431–5452.e20 (2024).

16. Natalini, J. G., Singh, S. & Segal, L. N. The dynamic lung microbiome in health and disease. Nat. Rev. Microbiol. 21, 222–235 (2023).

17. Huffnagle, G. B. & Dickson, R. P. The bacterial microbiota in inflammatory lung diseases. Clinical Immunology 159, 177–182 (2015).

18. Pattaroni, C. et al. Early-Life Formation of the Microbial and Immunological Environment of the Human Airways. Cell Host Microbe 24, 857–865.e4 (2018).

19. Gleeson, K., Maxwell, S. L. & Eggli, D. F. Quantitative aspiration during sleep in normal subjects. Chest 111, 1266–1272 (1997).

20. Sulaiman, I. et al. Functional lower airways genomic profiling of the microbiome to capture active microbial metabolism. Eur. Respir. J. 58, (2021).

21. Wu, B. G. et al. Episodic Aspiration with Oral Commensals Induces a MyD88-dependent, Pulmonary T-Helper Cell Type 17 Response that Mitigates Susceptibility to Streptococcus pneumoniae. Am. J. Respir. Crit. Care Med. 203, 1099–1111 (2021).

22. Horn, K. J., Schopper, M. A., Drigot, Z. G. & Clark, S. E. Airway Prevotella promote TLR2-dependent neutrophil activation and rapid clearance of Streptococcus pneumoniae from the lung. Nat. Commun. 13, 3321 (2022).

23. Kelly, M. S. et al. Role of the upper airway microbiota in respiratory virus and bacterial pathobiont dynamics in the first year of life. Nat. Commun. 16, 5195 (2025).

24. Chanderraj, R. et al. In critically ill patients, anti-anaerobic antibiotics increase risk of adverse clinical outcomes. Eur. Respir. J. 61, (2023).

25. Kitsios, G. D. et al. The upper and lower respiratory tract microbiome in severe aspiration pneumonia. iScience 26, 106832 (2023).

26. Kullberg, R. F. J., Schinkel, M. & Wiersinga, W. J. Empiric anti-anaerobic antibiotics are associated with adverse clinical outcomes in emergency department patients. Eur. Respir. J. 61, (2023).

27. Sirota, S. B. et al. Global burden of lower respiratory infections and aetiologies, 1990-2023: a systematic analysis for the Global Burden of Disease Study 2023. Lancet Infect. Dis. 26, (2026).

28. O’Brien, K. L. et al. Burden of disease caused by Streptococcus pneumoniae in children younger than 5 years: global estimates. Lancet 374, 893–902 (2009).

29. Li, L., Li, Y., Wang, Y., Fu, S. & Song, C. Differences in immune function and cytokine levels among children in different age groups with severe community-acquired pneumonia. Sci. Rep. 15, (2025).

30. Gopal, V. et al. Does Prior Respiratory Viral Infection Provide Cross-Protection Against Subsequent Respiratory Viral Infections? A Systematic Review and Meta-Analysis. Viruses 16, (2024).

31. Ohuma, E. O. et al. The natural history of respiratory syncytial virus in a birth cohort: the influence of age and previous infection on reinfection and disease. Am. J. Epidemiol. 176, 794–802 (2012).

32. Fernandes, C. D. et al. Host Inflammatory Biomarkers of Disease Severity in Pediatric Community-Acquired Pneumonia: A Systematic Review and Meta-analysis. Open Forum Infect. Dis. 6, (2019).

33. Chisti, M. J., Tebruegge, M., La Vincente, S., Graham, S. M. & Duke, T. Pneumonia in severely malnourished children in developing countries - mortality risk, aetiology and validity of WHO clinical signs: a systematic review. Trop. Med. Int. Health 14, 1173–1189 (2009).

34. Butters, C., Phuong, L. K., Cole, T. & Gwee, A. Prevalence of Immunodeficiency in Children With Invasive Pneumococcal Disease in the Pneumococcal Vaccine Era: A Systematic Review. JAMA Pediatr. 173, 1084–1094 (2019).

35. Tapia, M. D. et al. The Etiology of Childhood Pneumonia in Mali: Findings From the Pneumonia Etiology Research for Child Health (PERCH) Study. Pediatr. Infect. Dis. J. 40, S18 (2021).

36. Lloyd-Price, J. et al. Strains, functions and dynamics in the expanded Human Microbiome Project. Nature 550, 61 (2017).

37. Salter, S. J. et al. Reagent and laboratory contamination can critically impact sequence-based microbiome analyses. BMC Biol. 12, 87-(2014).

38. Piro, V. C. & Renard, B. Y. Contamination detection and microbiome exploration with GRIMER. Gigascience 12, 1–13 (2022).

39. Laurence, M., Hatzis, C. & Brash, D. E. Common Contaminants in Next-Generation Sequencing That Hinder Discovery of Low-Abundance Microbes. PLoS One 9, e97876 (2014).

40. Deloria-Knoll, M. et al. Identification and selection of cases and controls in the Pneumonia Etiology Research for Child Health project. Clinical Infectious Diseases 54 **Suppl 2**, S117–23 (2012).

41. Xu, R. et al. Longitudinal Profiling of the Human Milk Microbiome from Birth to 12 Months Reveals Overall Stability and Selective Taxa-Level Variation. Microorganisms 13, 1830 (2025).

42. Cheng, A. & Holland, S. M. Anti-cytokine autoantibodies: mechanistic insights and disease associations. Nat. Rev. Immunol. 24, 161–177 (2024).

43. Bastard, P. et al. Autoantibodies against type I IFNs in patients with life-threatening COVID-19. Science 370, (2020).

44. Feng, A., et al. Autoantibodies are highly prevalent in non-SARS-CoV-2 respiratory infections and critical illness. JCI Insight 8, (2023).

45. Chang, S. E. et al. New-onset IgG autoantibodies in hospitalized patients with COVID-19. Nat. Commun. 12, (2021).

46. Bastard, P. et al. Higher COVID-19 pneumonia risk associated with anti-IFN-α than with anti-IFN-ω auto-Abs in children. J. Exp. Med. 221, (2024).

47. Bastard, P. et al. Autoantibodies against type I IFNs in patients with life-threatening COVID-19. Science 370, (2020).

48. Chang, S. E. et al. New-onset IgG autoantibodies in hospitalized patients with COVID-19. Nat. Commun. 12, (2021).

49. Feng, A., et al. Autoantibodies are highly prevalent in non-SARS-CoV-2 respiratory infections and critical illness. JCI Insight 8, (2023).

50. Martín, V., Mañes-Lázaro, R., Rodríguez, J. M. & Maldonado-Barragán, A. Streptococcus lactarius sp. nov., isolated from breast milk of healthy women. Int. J. Syst. Evol. Microbiol. 61, 1048–1052 (2011).

51. Gehrig, J. L. et al. Effects of microbiota-directed foods in gnotobiotic animals and undernourished children. Science 365, (2019).

52. Raman, A. S. et al. A sparse covarying unit that describes healthy and impaired human gut microbiota development. Science (1979). 365, (2019).

53. Kelly, M. S. et al. Non-diphtheriae Corynebacterium species are associated with decreased risk of pneumococcal colonization during infancy. ISME J. 16, 655–665 (2022).

54. Stubbendieck, R. M., Hurst, J. H. & Kelly, M. S. Dolosigranulum pigrum: A promising nasal probiotic candidate. PLoS Pathog. 20, e1011955 (2024).

55. Sorbara, M. T. & Pamer, E. G. Interbacterial mechanisms of colonization resistance and the strategies pathogens use to overcome them. Mucosal Immunol. 12, 1–9 (2019).

56. Dong, Q. et al. Protection against Clostridioides difficile disease by a naturally avirulent strain. Cell Host Microbe 33, 59–70.e4 (2025).

57. Spragge, F. et al. Microbiome diversity protects against pathogens by nutrient blocking. Science (1979). 382, eadj3502 (2023).

58. Atto, B., Kunde, D., Gell, D. A. & Tristram, S. Oropharyngeal Carriage of hpl-Containing Haemophilus haemolyticus Predicts Lower Prevalence and Density of NTHi Colonisation in Healthy Adults. Pathogens 10, (2021).

59. Frivold, C. et al. Correlates of risk of respiratory syncytial virus disease: a prospective cohort study. Nat. Commun. 16, (2025).

60. Ohuma, E. O. et al. The natural history of respiratory syncytial virus in a birth cohort: the influence of age and previous infection on reinfection and disease. Am. J. Epidemiol. 176, 794–802 (2012).

61. Gopal, V. et al. Does Prior Respiratory Viral Infection Provide Cross-Protection Against Subsequent Respiratory Viral Infections? A Systematic Review and Meta-Analysis. Viruses 16, (2024).

62. Piret, J. & Boivin, G. The impact of trained immunity in respiratory viral infections. Rev. Med. Virol. 34, (2024).

63. Noma, K., Asano, T., Taniguchi, M., Ashihara, K. & Okada, S. Anti-cytokine autoantibodies in human susceptibility to infectious diseases: insights from Inborn errors of immunity. Immunol. Med. 48, 124–140 (2025).

64. Burbelo, P. D. et al. Rapid induction of autoantibodies during ARDS and septic shock. J. Transl. Med. 8, (2010).

65. Shaw, E. R. et al. Temporal Dynamics of Anti-Type 1 Interferon Autoantibodies in Patients With Coronavirus Disease 2019. Clin. Infect. Dis. 75, E1192–E1194 (2022).

66. Ruhamyankaka, E. et al. ClinEpiDB: an open-access clinical epidemiology database resource encouraging online exploration of complex studies. Gates Open Res. 3, 1661 (2019).

67. Crawley, J. et al. Standardization of Clinical Assessment and Sample Collection Across All PERCH Study Sites. Clin. Infect. Dis. 64, S228 (2017).

68. Clarke, E. L. et al. Sunbeam: an extensible pipeline for analyzing metagenomic sequencing experiments. Microbiome 7, 46 (2019).

69. Wood, D. E. & Salzberg, S. L. Kraken: Ultrafast metagenomic sequence classification using exact alignments. Genome Biol. 15, R46-(2014).

70. Ogata, H. et al. KEGG: Kyoto Encyclopedia of Genes and Genomes. Nucleic Acids Res. 27, 29–34 (1999).

71. Chen, L. et al. VFDB: a reference database for bacterial virulence factors. Nucleic Acids Res. 33, D325 (2004).

72. Buchfink, B., Reuter, K. & Drost, H. G. Sensitive protein alignments at tree-of-life scale using DIAMOND. Nat. Methods 18, 366–368 (2021).

73. Chang, S. E. et al. New-onset IgG autoantibodies in hospitalized patients with COVID-19. Nature Communications 2021 12:1 12, 5417-(2021).

74. Jiang, Y., Biecek, P., Paluszyńska, O., agasitko & KasiaKobylinska. ModelOriented/randomForestExplainer: CRAN release 0.10.1. 10.5281/ZENODO.3941250 doi:10.5281/ZENODO.3941250.

75. Oliveira, F. S. et al. MicrobiomeDB: a systems biology platform for integrating, mining and analyzing microbiome experiments. Nucleic Acids Res. 46, D684–D691 (2018).

