## Supplemental Figure 1 for "Microbial and immune determinants of disease severity and death in pediatric pneumonia"

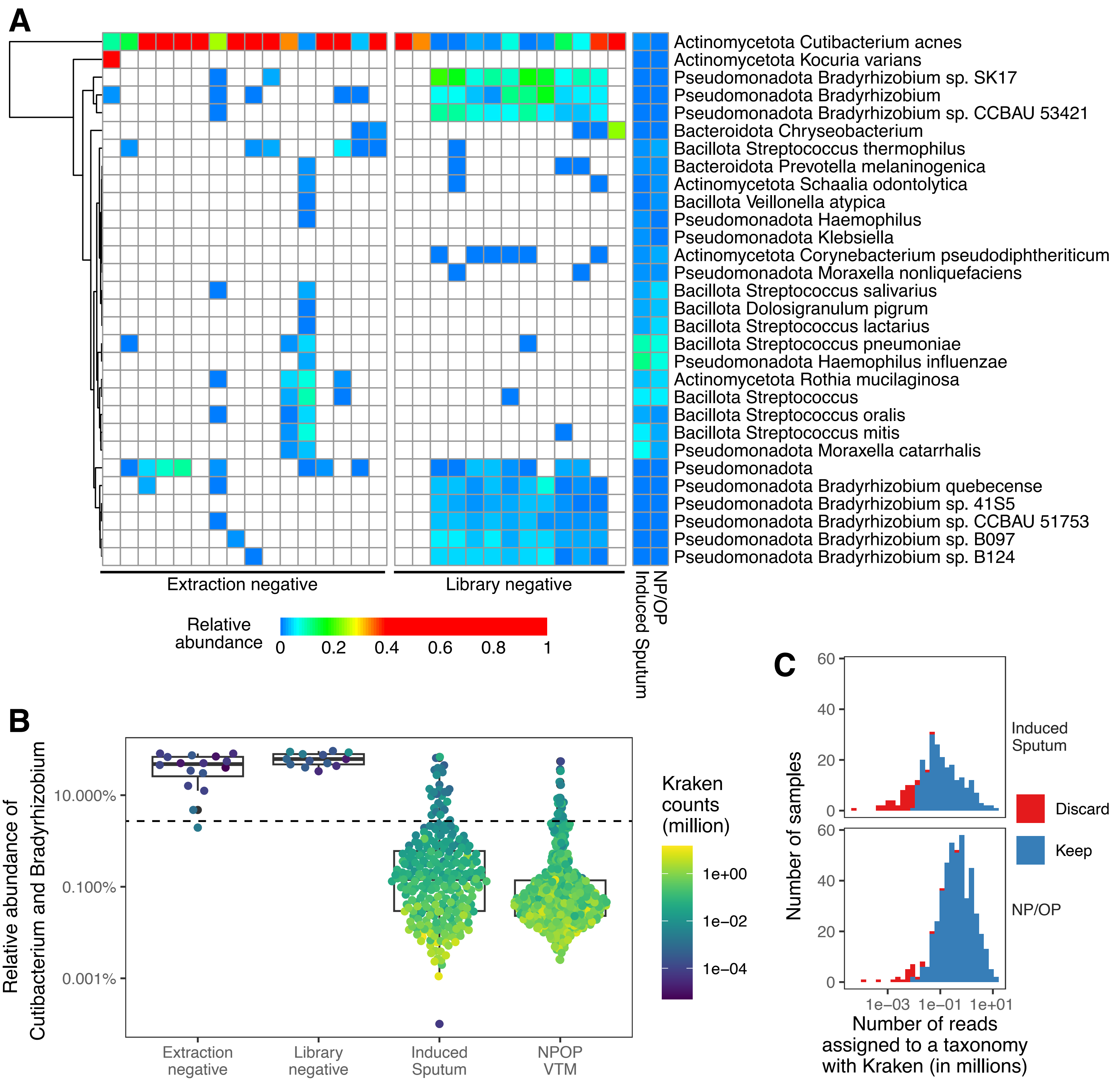


**Supplemental Figure 1: Quality control filtering of PERCH microbiome samples. A)** Heatmap of extraction and library negative controls and average relative abundance of NP/OP and induced sputum samples across the cohort. Only the taxa with 1.5% relative abundance in at least one sample type are shown. **B)** Relative abundance of the sum of Cutibacterium and Bradyrhizobium genera in all sample types. The dashed line represents the 2 standard deviations below the mean of control samples which was used as a threshold to exclude samples from NP/OP and sputum samples. **C)** The number of reads assigned to taxonomy using Kraken in the cohort. The number of samples discarded based on the threshold established in B and 10,000 Kraken assigned read count are shown in red.
