## Supplemental Figure 2 for "Microbial and immune determinants of disease severity and death in pediatric pneumonia"

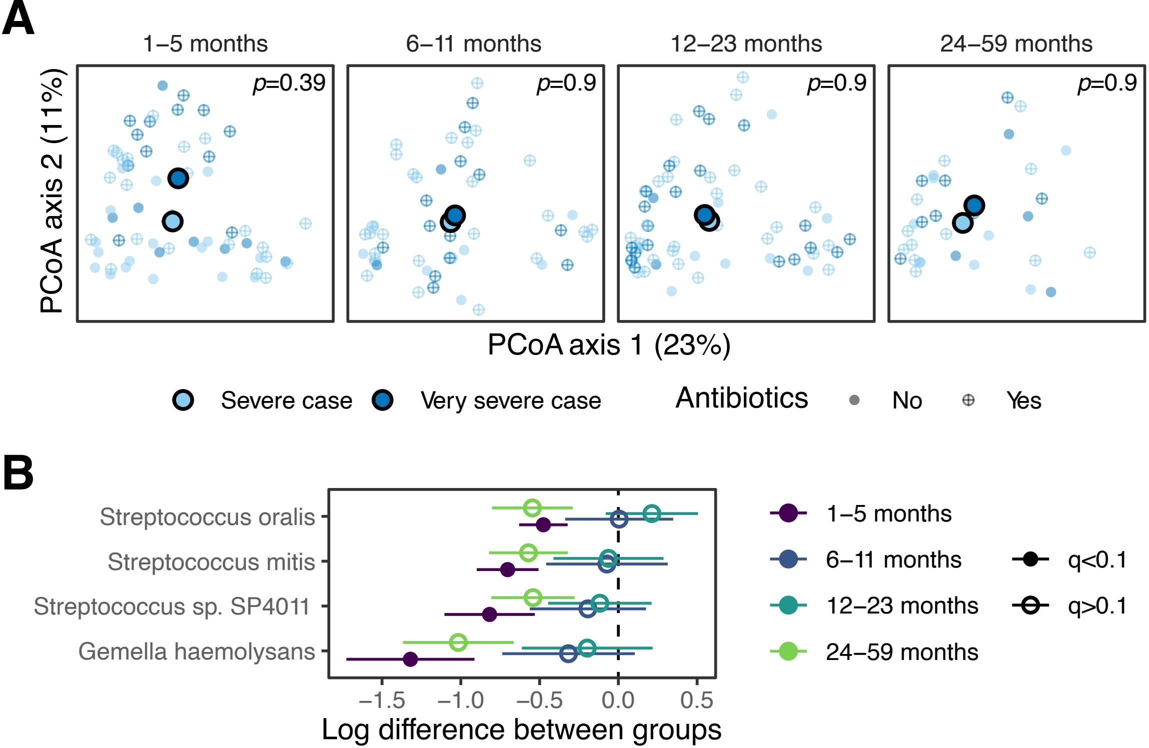


**Supplemental Figure 2: Disease severity- and age-dependent changes in microbial composition of induced sputum. A)** Principal component analysis of taxonomic composition of induced sputum samples. The axis labels represent the percent variation captured by the axes. PERMANOVA test was conducted to check if the centroids of the severe and very severe pneumonia groups could be distinguished from each other. **B)** Log difference of taxon relative abundance between severe and very severe groups. Only the taxa with q<0.1 in at least one comparison is shown.
