## Supplemental Figure 3 for "Microbial and immune determinants of disease severity and death in pediatric pneumonia"

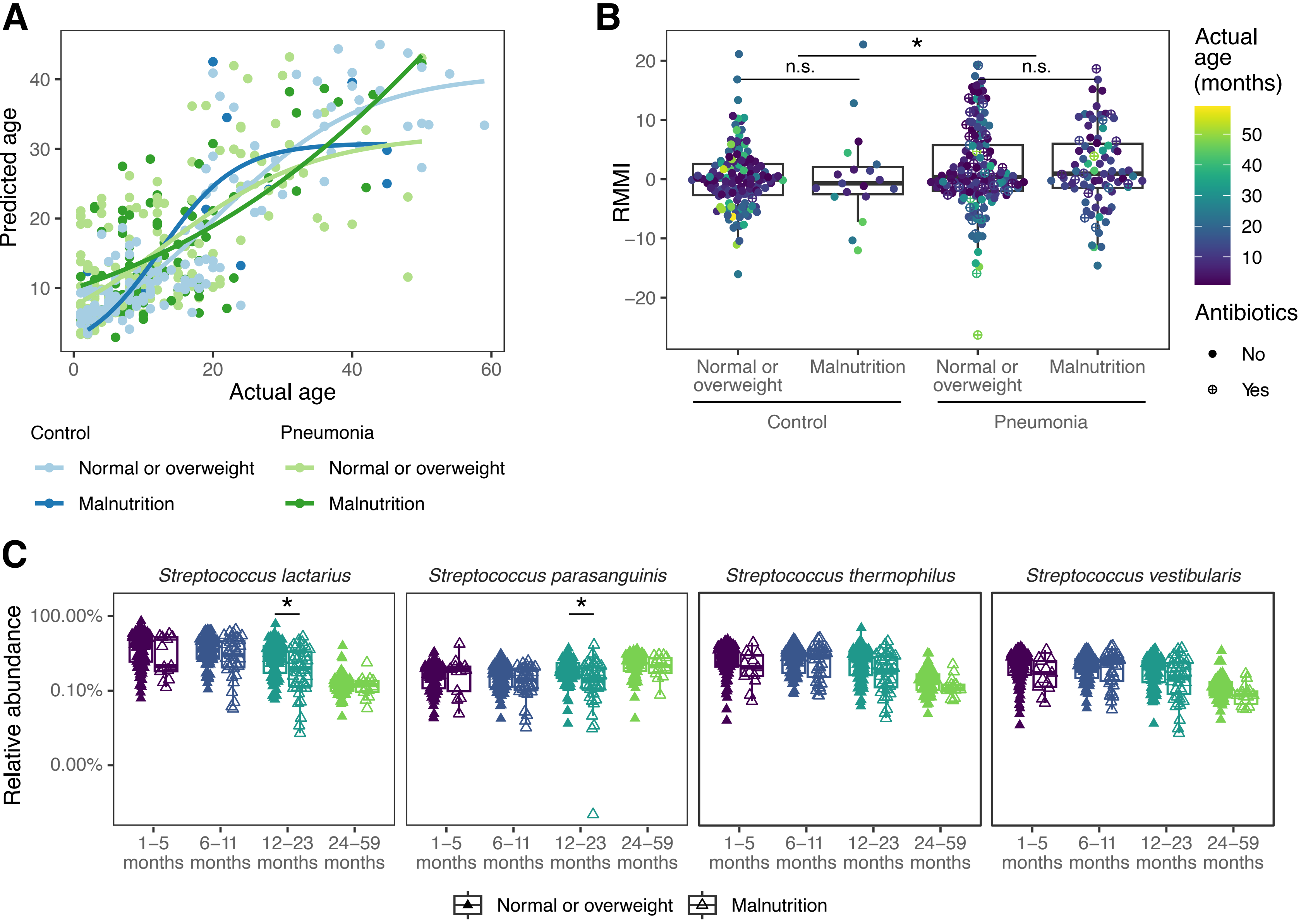


**Supplemental Figure 3: Malnutrition is not a major factor in age-dependent maturation of the upper respiratory tract (URT) microbiome. A)** Predicted age of pneumonia and control subjects with other without malnutrition using the random forest model built in Figure 1H. Line represents the self-starting logistic curve. **B)** relative maturity index calculated using a random forest model built with the ages as outcome and the relative abundance of taxa with at least 0.5% mean across the NP/OP samples of control and pneumonia participants (malnourished and non-malnourished) as predictors. **C)** Relative abundance of breastfeeding associated bacteria in malnourished or not malnourished subjects.
