## Supplemental Figure 4 for "Microbial and immune determinants of disease severity and death in pediatric pneumonia"

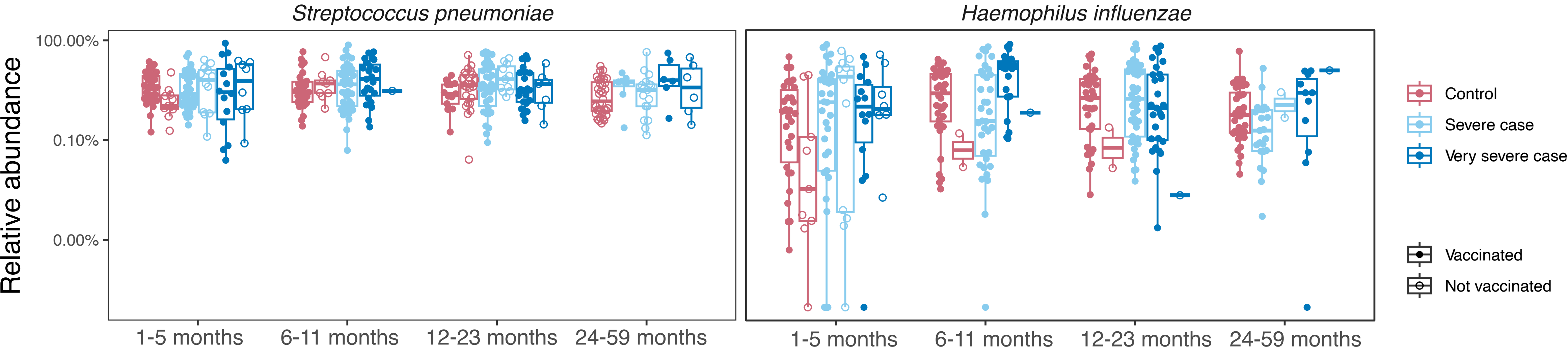


**Supplemental Figure 4: *Streptococcus pneumoniae* and *Haemophilus influenzae* levels in the URT are not impacted by vaccination.** Box and whisker plots showing relative abundance of **A)** *Streptococcus pneumoniae* and **B)** *Haemophilus influenzae* in the URT by age group, disease status, and vaccination status.
